# Physical and psychosocial outcomes in children with limb loss 2-years after the 2023 Kahramanmaraş-Türkiye earthquakes

**DOI:** 10.64898/2026.11.13.26367810

**Authors:** Fraje Watson, Aylin Sariyildiz, Ezgi Nur Sayici, Özge Metin, Bugra Kundakci, Anthony M. J. Bull, Angela E. Kedgley, Sibel Basaran

**Author notes:** joint first authorship (Fraje Watson, Aylin Sariyildiz).

## Abstract

**Introduction:** The 2023 Kahramanmaraş-Türkiye earthquakes resulted in many paediatric amputations. While prosthetic provision following disasters has been described, little is known about longer-term physical and psychosocial outcomes in children with earthquake-related limb loss.

**Aim:** Evaluate health-related quality of life, prosthesis experience, pain, and function approximately two years post-amputation.

**Method:** Retrospective analysis was conducted of children with earthquake-related limb loss who attend the XXX Center. Socio-demographic and amputation-related data, psychological status, outcome scores for pain, the Paediatric Quality of Life Inventory (PedsQL) and the Trinity Amputation and Prosthesis Experience Scales (TAPES) were included.

**Results:** Fifty-six children (50% female; 13.7±5.0 years old) were included. Children had unilateral upper limb loss (n=15), unilateral lower limb loss (n=31), bilateral lower limb loss (n=7), or combined upper and lower limb loss (n=3). Data were obtained 2.3±0.4 years following amputation. Median child-reported PedsQL scores were 72.3/100.0, with high social functioning (85.0/100.0) but lower emotional functioning (60.0/100.0). No differences in PedsQL outcomes were found between limb loss groups. Children rated their physical and emotional quality of life higher than carers rated them. TAPES median scores were 50.0/75.0 for psychosocial adjustment, 16.0/36.0 for activity restriction, and 34.5/45.0 for prosthetic satisfaction. Children with unilateral lower limb loss demonstrated superior adjustment and prosthetic satisfaction than children with unilateral upper limb or bilateral lower limb loss (p<0.05).

**Conclusions:** Children demonstrated favourable physical, social, and school-related quality of life 2.3 years after the earthquakes. However, emotional wellbeing remained affected, highlighting the importance of long-term mental health support alongside prosthetic and rehabilitation services.

## Introduction

Earthquakes often lead to complex, highly mechanical orthopaedic injuries to the lower limbs alongside complex soft tissue complications ^1^. Limb amputation can be required where injuries are too complex for salvage, or where crush syndrome has caused life-threatening muscle necrosis ^1^. In addition to immediate medical needs, earthquakes often bring widespread societal challenges across healthcare, education, jobs, and accommodation that further compound the effects and result in long-lasting repercussions ^2^. Children are particularly susceptible to both primary injuries and societal challenges due to their reliance on adults for their care.

Psychiatric outcomes of children following the 2023 Kahramanmaraş-Türkiye earthquakes has shown high rates of anxiety, depression, acute stress disorder, post-traumatic stress disorder and sleep problems ^3-5^. These outcomes were more prevalent in children who experienced bereavement, damage to their home, witnessed other people trapped in buildings, and sustained a physical injury. In one study, increasing family support was associated with decreasing risk of post-traumatic stress disorder ^3^.

Physical and psychosocial outcome data of children with limb loss following an earthquake are limited and there are few studies that examine longitudinal outcomes. In some cases, follow-up has taken place during early adulthood; however, results are rarely dichotomised to consider those who were children at the point of amputation ^6,7^. One study of 16 young adults 7-years post-unilateral transfemoral limb loss, who were children at the time of the 2008 Sichuan earthquake, found they had better psychosocial adjustment to their prosthesis and better overall physical health than those who were adults and sustained limb loss at the time of the earthquake ^8^. Whilst an important observation, this study did not include children who were less than 10 years old at the time of the earthquake or children with bilateral, upper or combination limb loss, nor did it provide evidence of satisfaction and quality of life during childhood and adolescence.

Health-related quality of life has been described in children with limb loss. A cohort of 15 children with predominantly unilateral, congenital lower limb loss in the UK had mean health-related quality of life scores of 67.6/100.0 ^9^, in comparison with 83.9/100.0 for children in the UK without chronic health conditions ^10^, where higher scores were better. Scores were similar to the cohort of 16 Cambodian children in the same study with predominantly unilateral traumatic lower limb loss who scored 67.8/100 ^9^. A cohort of 42 Turkish children with upper and lower limb loss for congenital and traumatic reasons reported better health-related quality of life using the same questionnaire (mean: 75.5/100.0) ^11^. Whilst these populations contain children with traumatic limb loss, these are more commonly due to road traffic accidents rather than natural disasters or earthquakes where injury epidemiology and environmental conditions are unique ^1^.

Primary information on 102 children treated by the Child Wellness Practice and Research Centre (CWC) following the 2023 Kahramanmaraş-Türkiye earthquakes has been described^12^. Our objective was to report physical and psychosocial outcomes of the same cohort of children up to three years after the earthquakes.

## Materials and methods

### Ethical approval

Ethical approval was granted by the Clinical Research Ethics Committee of Cukurova University Faculty of Medicine (date: 13 March 2026; approval number: 164-40). Informed consent was obtained from a parent/carer.

### Study participants

Participants were all children who sustained limb loss in the 2023 Kahramanmaraş-Türkiye Earthquakes and attended CWC. A convenience sample of children who completed a set of three questionnaires at a minimum of one time point were included. For children with multiple responses, those collected at the longest time since the earthquake was included.

### Patient-reported outcome measures

#### PedsQL-Pain

The Paediatric Pain Questionnaire (PedsQL-Pain) is a reliable and valid assessment of pain in children ^13^, which was translated into Turkish. Children and parent/carer proxies scored pain on the day and pain in the seven days prior along a visual analog scale labelled from “it doesn’t hurt, no discomfort, no pain” to “very uncomfortable, severe pain”.

#### PedsQL-QoL

The Paediatric Quality of Life Inventory™ (PedsQL-QoL) is a reliable and valid assessment of health-related quality of life in healthy children ^14^. PedsQL consists of 23-items across four scales; physical, emotional, social, and school, scored on a five-point Likert scale, scored by both the child and parent/carer. Scores are computed as 0-100, where a higher score indicates a better outcome. PedsQL is valid and reliable when translated into Turkish ^15-17^.

#### TAPES

The Trinity Amputation and Prosthesis Experience Scales (TAPES) comprises three domains each with three subdomains; psychosocial adjustment (general, social, adjustment to limitation), activity restriction (functional, social, athletic), and prosthetic satisfaction (functional, aesthetic, weight) of lower limb amputees ^18^. Scales are measured using three- and five-point scales. Psychological adjustment subdomains are scored 5-25 (total: 15-75), where a higher score indicates better adjustment. The activity restriction scale is scored 4-12 (total: 12-36), where a higher score indicates more activity restriction. The functional and aesthetic subdomains of prosthetic satisfaction are scored 4-20, and weight is scored 1-5 (total: 9-45), where a higher score indicates better adjustment. TAPES is reliable and valid in Turkish ^19^. TAPES was used to assess all children, but those with upper limb (UL) loss only completed the psychosocial adjustment and satisfaction sections.

#### CAPP-FSI

The Child Amputee Prosthetics Project – Functional Status Inventory (CAPP-FSI) is a reliable and valid assessment designed to measure the functional ability of children with upper and lower limb difference ^20^. Children with upper limb loss scored their frequency of conducting 34 activities both with and without their prosthesis on a five-point scale, scoring 0-144 where a higher score is better. Children with lower limb loss scored their frequency of conducting 6 activities both with and without their prosthesis on the same five-point scale scoring 0-24. The CAPP-FSI has been translated into Turkish ^21^.

### Socioeconomic outcomes

Monthly family income was categorised in respect to the hunger line and the poverty line in the month the information was collected as published by the Confederation of Turkish Trade Unions, TÜRK-İŞ ^22^.

### Statistical analysis

Clinical and outcome datapoints were matched to psychiatric data collected within three months of each other. For each child, the timepoint with the longest complete follow-up was kept. Normality was determined by visual inspection of histograms and Q-Q plots. Normally distributed data were reported as mean (standard deviation; SD) and non-normally distributed data reported as median (interquartile range; IQR). When continuous, non-parametric variables were compared between three amputation groups, a Kruskal-Wallis test was used. Where a difference was found, post-hoc comparison was calculated with Bonferroni correction. When continuous, non-parametric variables were compared between paired units (e.g., child and parent/carer) a Wilcoxon signed-rank test was used.

## Results

### Study participants

Fifty-six children were included in the study. Children were 50% (n=28) female with a mean age of 11.6 years (SD: 4.8 years) at the earthquake, and 13.7 years (SD: 5.0 years) at follow-up. Children sustained unilateral upper limb loss (n=15), unilateral lower limb loss (n=31), bilateral lower limb loss (n=7), unilateral upper and unilateral lower limb loss (n=1), and unilateral upper and bilateral lower limb loss (n=2) (Table 1).

**Table 1:** Demographic information for the full cohort and unilateral upper limb (UL), unilateral lower limb (LL), bilateral LL, unilateral UL, unilateral LL, and unilateral UL, bilateral LL amputation groups.

|  | Full cohort | Unilateral UL | Unilateral LL | Bilateral LL | Unilateral UL, unilateral LL | Unilateral UL bilateral LL |
|---|---|---|---|---|---|---|
| n | 56 | 15 | 31 | 7 | 1<sup>%</sup> | 2<sup>%</sup> |
| Amputation level | - | SD: 1<br>TH: 10<br>TR: 2<br>Partial: 2 | HD: 1<br>TF: 5<br>KD: 8<br>TT: 13<br>Syme: 1<br>Chopart: 1<br>Partial: 2 | HD+TF: 1<br>TF+TF: 2<br>KD+TT: 1<br>TT+TT: 2<br>TT+partial: 1 | TF+partial: 1 | SD+TF+partial: 1<br>TH+TF+TT: 1 |
| Age at earthquake; yr | 11.3 (5.0) | 11.5 (5.7) | 11.5 (4.8) | 11.2 (5.7) | - | 8.3 (5.1) |
| Age at amputation; yr | 11.4 (5.0) | 11.5 (5.7) | 11.5 (4.8) | 11.2 (5.7) | - | 8.4 (5.0) |
| Age at follow-up; yr | 13.7 (5.0) | 13.7 (5.6) | 13.9 (4.8) | 13.4 (5.5) | - | 10.6 (5.5) |
| Amputation to follow-up; yr | 2.3 (0.4) | 2.2 (0.4) | 2.4 (0.4) | 2.2 (0.5) | - | 2.2 (0.5) |
| Earthquake to follow-up; yr | 2.4 (0.4) | 2.2 (0.4) | 2.4 (0.4) | 2.2 (0.5) | - | 2.5 (0.4) |
| Female; count (%) | 28 (50.0) | 7 (46.7) | 17 (54.8) | 2 (28.6) | - | - |
| Height; cm | 151.5 (20.8) | 153.0 (23.2) | 152.7 (19.1) | 148.4 (28.0) | 149.0 | 134.0 (25.5) |
| Weight (including prosthesis); kg | 50.8 (20.3) | 55.1 (24.1) | 51.0 (18.6) | 46.2 (24.4) | 43.8 | 40.3 (28.7) |
| Time under rubble; h | 43.1 (40.5) | 22.9 (29.1) | 46.3 (40.1) | 49.0 (31.3) | - | 64.0 (11.3) |
| Lost first-degree family member |  |  |  |  |  |  |
| Yes; count (%) | 42 (75.0) | 9 (60.0) | 24 (77.4) | 6 (85.7) | - | - |
| Number; median (IQR) | 1 (0.5-2.5) | 1 (0-3) | 2 (1-3) | 1 (1-2) | - | - |
| At least one parent died in the earthquakes; count (%) | 35 (62.5) | 9 (60.0) | 20 (64.5) | 3 (42.9) | - | - |
| Both parents died in the earthquakes; count (%) | 16 (28.6) | 3 (20.0) | 10 (32.3) | 1 (14.3) | - | - |
| Family amputation (yes); count (%) | 13 (23.2) | 4 (26.7) | 7 (22.6) | 2 (28.6) | - | - |
| Psychiatric diagnosis; count (%) |  |  |  |  |  |  |
| Yes, trauma-related | 27 (48.2) | 6 (40.0) | 15 (48.4) | 5 (71.4) | - | - |
| Yes, not trauma-related | 9 (16.1) | 2 (13.3) | 5 (16.1) | 2 (28.6) |  |  |
| No | 16 (28.6) | 5 (33.3) | 9 (29.0) | 0 (0.0) |  |  |
| Unknown | 4 (7.1) | 2 (13.3) | 2 (6.5) | 0 (0.0) |  |  |
| Trauma-related psychiatric diagnoses; count (%) |  |  |  |  |  |  |
| Depression | 5 (8.9) | 1 (6.7) | 1 (3.2) | 3 (42.9) | - | - |
| Anxiety | 6 (10.7) | 0 (0.0) | 3 (9.7) | 2 (28.6) |  |  |
| PTSD | 14 (25.0) | 3 (20.0) | 9 (29.0) | 2 (28.6) |  |  |
| Other | 5 (8.9) | 2 (13.3) | 2 (6.5) | 1 (14.3) |  |  |
| Housing; count (%) |  |  |  |  |  |  |
| Shelter | 15 (26.8) | 6 (40.0) | 6 (19.4) | 2 (28.6) | - | - |
| Pre-fab | 1 (1.8) | 0 (0.0) | 1 (3.2) | 0 (0.0) |  |  |
| House | 38 (67.9) | 8 (53.3) | 23 (74.2) | 5 (71.4) |  |  |
| Unknown | 2 (3.6) | 1 (6.7) | 1 (3.2) | 0 (0.0) |  |  |
| Family income; count (%) |  |  |  |  |  |  |
| Below hunger line* | 25 (44.6) | 6 (40.0) | 16 (51.6) | 2 (28.6) | - | - |
| Below poverty line* | 41 (73.2) | 8 (53.3) | 9 (29.0) | 6 (85.7) |  |  |
| Unknown | 12 (21.4) | 4 (26.7) | 6 (19.4) | 0 (0.0) |  |  |
| Attends school<sup>#</sup> (no); count (%) | 1 (1.8) | 1 (6.7) | 0 (0.0) | 0 (0.0) | - | - |

Unless otherwise stated, data are presented as mean (standard deviation). Non-parametric data is presented as median (interquartile range; IQR) and count data is presented as count followed by percentage (%).
<sup>%</sup>Information redacted to protect child's identity due to n=1 and n=2
<sup>&</sup>height of children with bilateral limb loss was measured according to age- and sex-specific Sitting Height to Stature Ratio
\*hunger line = minimum monthly cost of a healthy and adequate food alone for a family of four in Ankara; poverty line = minimum monthly cost of a healthy and adequate food and all other essential living expenses for a family of four in Ankara <sup>22</sup>
<sup>#</sup>For school age children in Türkiye (6-18-years old)
ADHD, Attention deficit hyperactivity disorder; PTSD, post-traumatic stress disorder; ODD, oppositional defiant disorder; OCD, obsessive-compulsive disorder; SD, shoulder disarticulation; TH, transhumeral; TR, transradial; HD, hip disarticulation; TF, transfemoral; KD, knee disarticulation; TT, transtibial

Forty-two (75.0%) children lost a first-degree family member in the earthquake, and 16 (28.6%) children lost both parents. Twenty-seven (48.2%) children were diagnosed with trauma-related psychiatric condition due to the earthquake. Forty-one (73.2%) children lived below the poverty line. For comparison, though slightly methodologically different, UNICEF reported that 32.7% of children in Türkiye lived in poverty in 2022 ^23^. Additional detail can be found in Table 1.

For children with upper limb loss; five used a cosmetic prosthesis and 10 used a myoelectric prosthesis. For children with lower limb loss; four used a SACH foot, 26 used a carbon foot and 15 used a dynamic foot. For children with lower limb loss using knee components; seven used a fixed/mechanical system and 15 used a hydraulic system. Further details on level of amputation and current prosthetic prescription are provided in Supplementary Table A.

Forty-six (82.1%) children used a prosthetic, and 37 (66.1%) of those reported daily use for a median of 10 hours a day (IQR: 6-13). From 50 children, 16 (32.0%) children reported stump pain, and 10 (20.4%) children reported phantom limb pain. One (1.8%) child with bilateral lower limb loss reported constant phantom limb pain. Additional detail can be found in Table 2.

**Table 2:** Demographic information for the full cohort and unilateral upper limb (UL), unilateral lower limb (LL), bilateral LL, unilateral UL, unilateral LL, and unilateral UL, bilateral LL amputation groups. Unless otherwise stated, data are presented as count (range).

|  | Full cohort | Unilateral UL | Unilateral LL | Bilateral LL | Unilateral UL, unilateral LL | Unilateral UL bilateral LL |
|---|---|---|---|---|---|---|
| n | 52 | 13 | 29 | 7 | 1 | 2 |
| Prosthesis use (yes); n (%) | 46 (88.5) | 10 (76.9) | 26 (89.7) | 7 (100.0) | 1 (100.0) | 2 (100.0) |
| Uses prosthesis daily (yes); n (%) | 37 | 6 (46.2) | 25 (86.2) | 5 (71.4) | 1 (100.0) | 1 (50.0) |
| Hours per day | 10 (6-13) | 6 (5-9) | 12 (8-15) | 9 (5-12) | 15 (-) | 10 (-) |
| Uses prosthesis weekly (yes); n (%) |  | 4 (30.8) | - | 2 (28.6) | - | 1 (50.0) |
| Hours per week |  | 3 (1.5-4.5) |  | 8 (6-10) |  | 8 (-) |
| Stump pain, n | 50 | 12 | 28 |  |  |  |
| Yes; count (%) | 16 (32.0) | 3 (25.0) | 7 (25.0) | 5 (71.4) | 0 (0.0) | 1 (50.0) |
| Frequency per week; n | 4 (3-7) | 3 (2-4) | 5.5 (3.5-7) | 3.5 (2.5-5.5) | - | 7 (-) |
| Pain duration; minutes | 30 (5-60) | 30.3 (0.5-60) | 10 (5-10) | 30 (30-60) | - | 40 (-) |
| Phantom sensation, n | 49 | 12 | 27 |  |  |  |
| Yes, n (%) | 17 (34.7) | 3 (25.0) | 11 (40.7) | 3 (42.9) | 0 (0.0) | 1 (50.0) |
| Phantom limb pain, n | 49 | 12 | 27 |  |  |  |
| Yes; n (%) | 10 (20.4) | 1 (8.3) | 8 (29.6) | 1 (14.3) | 0 (0.0) | 0 (0.0) |
| Frequency per week; n | 4 (3-7) | 7 (-) | 3.5 (3-5.5) | 7 (-) | - | - |
| Pain duration; minutes | 5 (3-5) | 5 (-) | 5 (2.5-12.5) | - | - | - |
| Pain constant (yes); n (%) | 1 | 0 (0.0) | 0 (0.0) | 1 (14.3) | - | - |

### Patient-reported outcome measures

#### PedsQL-Pain

PedsQL-Pain scores (Table 3) were available for 44 (78.6%) children and 37 (66.1%) carer proxies. Median pain score on the day was 0.3 (IQR: 0.0-2.7) for children and 0.8 (IQR: 0.0-3.4) scored by carer proxies. Median pain scores for the seven days prior were 2.4 (IQR: 0.0-5.9) for children and 2.5 (IQR: 0.0-4.0) scored by carer proxies.

**Table 3:**
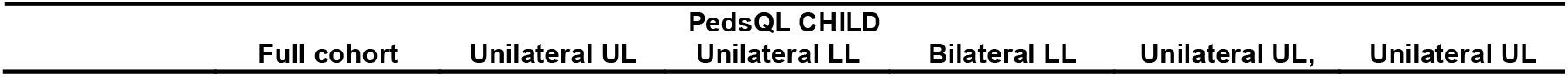
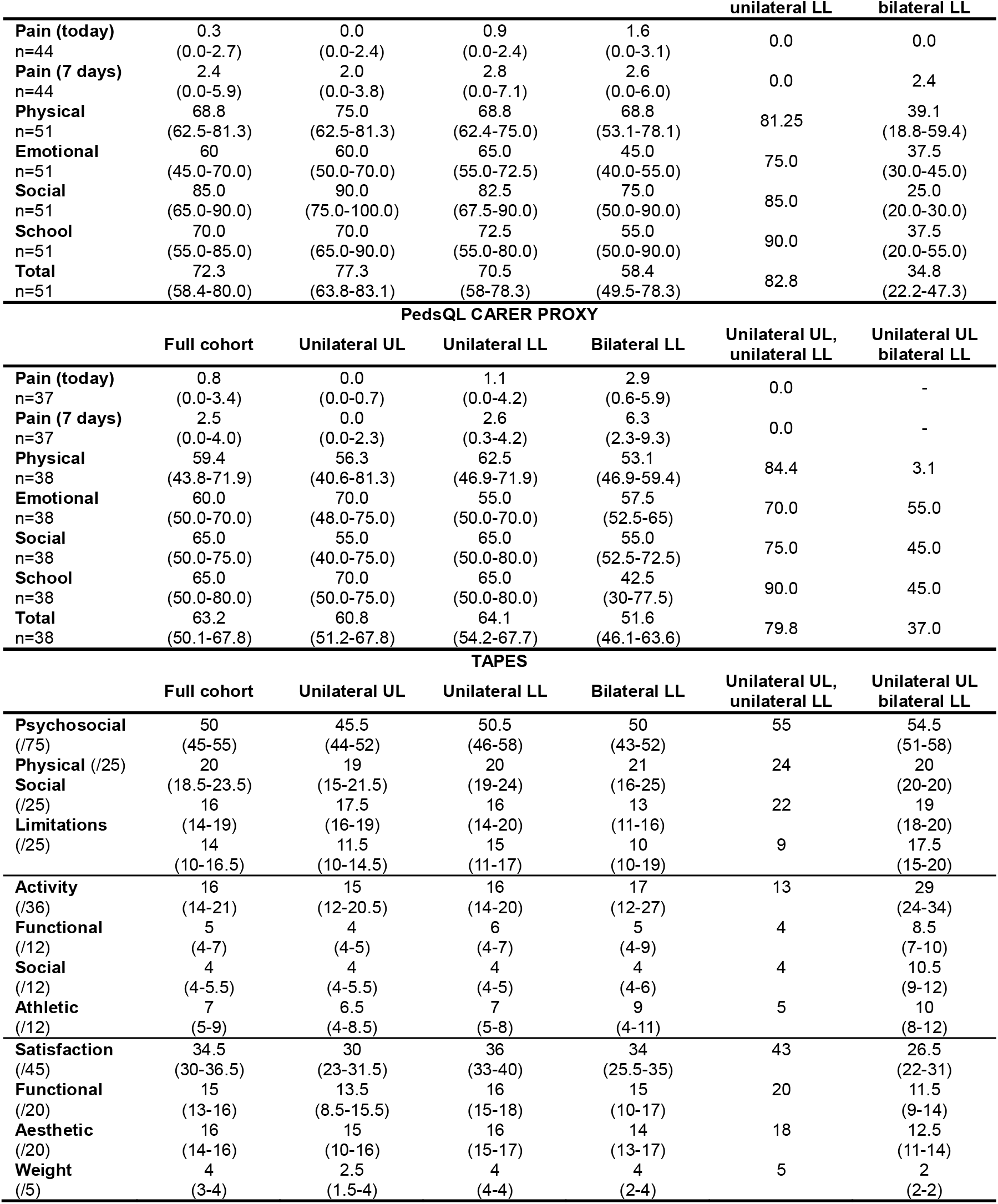
Paediatric Quality of Life Inventory (PedsQL) for pain on the day of the survey (today), pain over the seven days prior (7 days), PedsQL-Physical, PedsQL-Emotional, PedsQL-Social, PedsQL-School and PedsQL-Total scored by the children and carer proxies, and Trinity Amputation and Prosthesis Experience Scales (TAPES) scores for the full cohort and each sub-group. Data are presented as median (interquartile range).

| PedsQL CHILD |  |  |  |  |  |  |
|---|---|---|---|---|---|---|
|  | **Full cohort** | **Unilateral UL** | **Unilateral LL** | **Bilateral LL** | **Unilateral UL,<br>unilateral LL** | **Unilateral UL<br>bilateral LL** |
| **Pain (today)**<br>n=44 | 0.3<br>(0.0-2.7) | 0.0<br>(0.0-2.4) | 0.9<br>(0.0-2.4) | 1.6<br>(0.0-3.1) | 0.0 | 0.0 |
| **Pain (7 days)**<br>n=44 | 2.4<br>(0.0-5.9) | 2.0<br>(0.0-3.8) | 2.8<br>(0.0-7.1) | 2.6<br>(0.0-6.0) | 0.0 | 2.4 |
| **Physical**<br>n=51 | 68.8<br>(62.5-81.3) | 75.0<br>(62.5-81.3) | 68.8<br>(62.4-75.0) | 68.8<br>(53.1-78.1) | 81.25 | 39.1<br>(18.8-59.4) |
| **Emotional**<br>n=51 | 60<br>(45.0-70.0) | 60.0<br>(50.0-70.0) | 65.0<br>(55.0-72.5) | 45.0<br>(40.0-55.0) | 75.0 | 37.5<br>(30.0-45.0) |
| **Social**<br>n=51 | 85.0<br>(65.0-90.0) | 90.0<br>(75.0-100.0) | 82.5<br>(67.5-90.0) | 75.0<br>(50.0-90.0) | 85.0 | 25.0<br>(20.0-30.0) |
| **School**<br>n=51 | 70.0<br>(55.0-85.0) | 70.0<br>(65.0-90.0) | 72.5<br>(55.0-80.0) | 55.0<br>(50.0-90.0) | 90.0 | 37.5<br>(20.0-55.0) |
| **Total**<br>n=51 | 72.3<br>(58.4-80.0) | 77.3<br>(63.8-83.1) | 70.5<br>(58-78.3) | 58.4<br>(49.5-78.3) | 82.8 | 34.8<br>(22.2-47.3) |
| **PedsQL CARER PROXY** |  |  |  |  |  |  |
|  | **Full cohort** | **Unilateral UL** | **Unilateral LL** | **Bilateral LL** | **Unilateral UL,<br>unilateral LL** | **Unilateral UL<br>bilateral LL** |
| **Pain (today)**<br>n=37 | 0.8<br>(0.0-3.4) | 0.0<br>(0.0-0.7) | 1.1<br>(0.0-4.2) | 2.9<br>(0.6-5.9) | 0.0 | - |
| **Pain (7 days)**<br>n=37 | 2.5<br>(0.0-4.0) | 0.0<br>(0.0-2.3) | 2.6<br>(0.3-4.2) | 6.3<br>(2.3-9.3) | 0.0 | - |
| **Physical**<br>n=38 | 59.4<br>(43.8-71.9) | 56.3<br>(40.6-81.3) | 62.5<br>(46.9-71.9) | 53.1<br>(46.9-59.4) | 84.4 | 3.1 |
| **Emotional**<br>n=38 | 60.0<br>(50.0-70.0) | 70.0<br>(48.0-75.0) | 55.0<br>(50.0-70.0) | 57.5<br>(52.5-65) | 70.0 | 55.0 |
| **Social**<br>n=38 | 65.0<br>(50.0-75.0) | 55.0<br>(40.0-75.0) | 65.0<br>(50.0-80.0) | 55.0<br>(52.5-72.5) | 75.0 | 45.0 |
| **School**<br>n=38 | 65.0<br>(50.0-80.0) | 70.0<br>(50.0-75.0) | 65.0<br>(50.0-80.0) | 42.5<br>(30-77.5) | 90.0 | 45.0 |
| **Total**<br>n=38 | 63.2<br>(50.1-67.8) | 60.8<br>(51.2-67.8) | 64.1<br>(54.2-67.7) | 51.6<br>(46.1-63.6) | 79.8 | 37.0 |
| **TAPES** |  |  |  |  |  |  |
|  | **Full cohort** | **Unilateral UL** | **Unilateral LL** | **Bilateral LL** | **Unilateral UL,<br>unilateral LL** | **Unilateral UL<br>bilateral LL** |
| **Psychosocial**<br>(/75) | 50<br>(45-55) | 45.5<br>(44-52) | 50.5<br>(46-58) | 50<br>(43-52) | 55 | 54.5<br>(51-58) |
| **Physical** (/25)<br>**Social** | 20<br>(18.5-23.5) | 19<br>(15-21.5) | 20<br>(19-24) | 21<br>(16-25) | 24 | 20<br>(20-20) |
| (/25)<br>**Limitations** | 16<br>(14-19) | 17.5<br>(16-19) | 16<br>(14-20) | 13<br>(11-16) | 22 | 19<br>(18-20) |
| (/25) | 14<br>(10-16.5) | 11.5<br>(10-14.5) | 15<br>(11-17) | 10<br>(10-19) | 9 | 17.5<br>(15-20) |
| **Activity**<br>(/36) | 16<br>(14-21) | 15<br>(12-20.5) | 16<br>(14-20) | 17<br>(12-27) | 13 | 29<br>(24-34) |
| **Functional**<br>(/12) | 5<br>(4-7) | 4<br>(4-5) | 6<br>(4-7) | 5<br>(4-9) | 4 | 8.5<br>(7-10) |
| **Social**<br>(/12) | 4<br>(4-5.5) | 4<br>(4-5.5) | 4<br>(4-5) | 4<br>(4-6) | 4 | 10.5<br>(9-12) |
| **Athletic**<br>(/12) | 7<br>(5-9) | 6.5<br>(4-8.5) | 7<br>(5-8) | 9<br>(4-11) | 5 | 10<br>(8-12) |
| **Satisfaction**<br>(/45) | 34.5<br>(30-36.5) | 30<br>(23-31.5) | 36<br>(33-40) | 34<br>(25.5-35) | 43 | 26.5<br>(22-31) |
| **Functional**<br>(/20) | 15<br>(13-16) | 13.5<br>(8.5-15.5) | 16<br>(15-18) | 15<br>(10-17) | 20 | 11.5<br>(9-14) |
| **Aesthetic**<br>(/20) | 16<br>(14-16) | 15<br>(10-16) | 16<br>(15-17) | 14<br>(13-17) | 18 | 12.5<br>(11-14) |
| **Weight**<br>(/5) | 4<br>(3-4) | 2.5<br>(1.5-4) | 4<br>(4-4) | 4<br>(2-4) | 5 | 2<br>(2-2) |

Pain scores for today were not different between amputation groups for children or carer proxies. Pain scores for the seven days prior were not different between amputation groups for children but were different for carer proxies (*p*=0.02). Post-hoc comparisons showed that the carer proxy score of pain for the seven days prior was higher in both unilateral LL and bilateral LL groups compared to the unilateral UL group (*p*=0.05 and *p*=0.01). There was no significant difference between child and carer proxy scores for pain today or in the last seven days for the 34 child-carer proxy pairs.

#### PedsQL-QoL

PedsQL-QoL scores were available for 51 (92.7%) children and 38 (67.9%) carer proxies. Median PedsQL-Total scores were 72.3 (IQR: 58.4-80.0) according to the children and 63.2 (IQR: 50.1-67.8) according to the carer proxies. Full results are available in Table 3.

Neither PedsQL-Total, PedsQL-Physical, PedsQL-Emotional, PedsQL-Social nor PedsQL-School were different between unilateral UL, unilateral LL and bilateral LL groups for children (Figure 1) or carer proxy scores. For the 36 child-carer pairs with complete data, PedsQL-Total, PedsQL-Physical and PedsQL-Emotional scores were all higher for children than carer proxy scores (*p*=0.015, *p*<0.001, *p*<0.001). Paired PedsQL-Emotional and PedsQL-School scores were not different between children and carer proxies.

**Figure 1:**
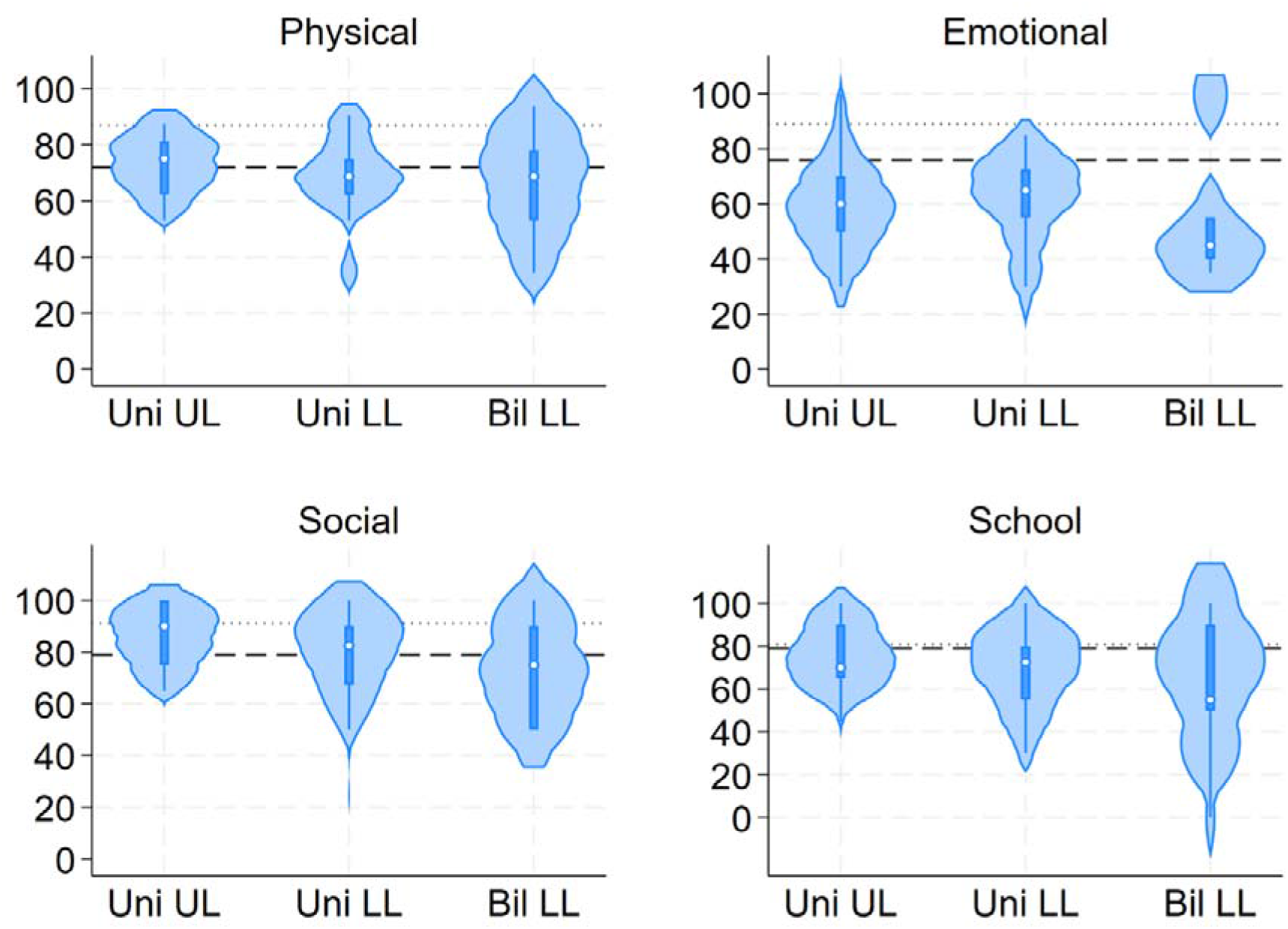
Violin plot for (A) Physical, (B) Emotional, (C) Social and (D) School components of the Paediatric Quality of Life (PedsQL) questionnaire completed by the children. The dotted line is the mean score for healthy Turkish children without limb loss ^24^. The dashed line is the mean score for Turkish children with non-earthquake related limb loss ^11^.

#### TAPES

TAPES scores were available for 52 (92.9%) participants (Table 3). Median psychosocial adjustment, activity restriction and prosthetic satisfaction scores were 50/75 (IQR:45-55), 16/36 (IQR: 14-21), and 34.5/75 (IQR: 30-36.5).

TAPES scores for psychosocial adjustment and prosthetic satisfaction were different between amputation groups (*p*=0.041 and *p*=0.001). Post-hoc comparisons showed that the unilateral LL group had higher psychosocial adjustment and prosthetic satisfaction scores than the unilateral UL (*p*=0.01 and *p*<0.001) and bilateral LL groups (*p*=0.05 and *p*=0.03). TAPES scores between Unilateral LL and Bilateral LL groups were not different for activity restriction.

#### CAPP-FSI

CAPP-FSI scores were available for 13 participants in the unilateral UL group. Median CAPP-FSI scores were 26 (IQR: 6-62) and 73 (IQR: 63-77) with and without their prosthetic. CAPP-FSI scores were available for 35 participants in the unilateral LL and bilateral LL groups. Median CAPP-FSI scores were 24 (IQR: 22-24) and 6 (IQR: 2-13) with and without their prosthetic.

## Discussion

These results demonstrate similar health-related quality of life approximately two years post-amputation of Turkish children who sustained unilateral UL, unilateral LL and bilateral LL loss following the 2023 Kahramanmaraş-Türkiye earthquakes. Though not compared statistically, there is evidence to suggest health-related quality of life is lower for children with triple amputation. Children received high-quality prosthetics and rehabilitation within a purpose-built centre that also supported them socially. The lasting psychosocial effects of the earthquake trauma itself and earthquake-related disruption of their family unit, school and housing are important factors in outcomes for these children.

PedsQL-Physical scores from children with unilateral upper limb loss were higher than, and scores from children with unilateral and bilateral lower limb loss were similar to those from a previously described cohort of Turkish children with non-earthquake-related limb loss ^11^. Scores across all groups were not as high as previously described in a comparison cohort of typically developing Turkish children ^24^. Similar physical health outcomes to other Turkish children with limb loss are an excellent result for children injured in the earthquakes because they were injured and provided with emergency treatment in a disaster zone, compared to other children who were treated within a functional healthcare system. However, the children in our cohort were treated within a dedicated rehabilitation centre, which may not be the case for all children with limb loss in Türkiye. Likely contributing to these good physical outcomes is the fact that children had generally low average pain scores, though a few children with unilateral and bilateral limb loss reported higher levels of pain in the seven days prior. The pain scores reported by our cohort of Turkish children are lower than those reported by children with limb loss from the UK and Cambodia ^9^.

Children in our cohort received high-quality prosthetics. However, TAPES scores in our cohort were all worse than outcomes from both Turkish children with non-earthquake-related limb loss and adults who sustained unilateral transfemoral amputation as children in the 2008 Sichuan earthquake ^8^. In Turkish children, this could represent the passage of time because the non-earthquake affected children were about 8-years post-amputation and half the cohort had congenital limb loss. In the Sichuan cohort, this could indicate some mismatch between expectation and reality, particularly as our cohort were still children themselves. In the UK, there was a distinct increase in visibility and public perception of people with limb loss following the 2012 London Paralympic Games ^25^. A positive change in public perception of people with limb loss between earthquakes occurring in 2008 and 2023 might contribute to increased expectations.

The CAPP-FSI demonstrated functional improvements in children with lower limb loss. However, in children with upper limb loss, scores were worse when using a prosthesis compared to without using a prosthesis. A similar pattern was seen in previous work in Turkish children with upper limb loss ^21^. The authors hypothesised that children had developed effective compensatory strategies, had basic prosthetics with limited functionality, and that CAPP-FSI rewarded task completion over prosthetic use. Furthermore, in our cohort of children, the CAPP-FSI was completed by children, whereas the original validation ^20^ and elsewhere ^21^, it is parent-reported or administered by an adult.

PedsQL-Social scores across all amputation groups were similar or higher than a previously described cohort of Turkish children with non-earthquake-related limb loss ^11^. In fact, scores were almost as high as previously described in a comparison cohort of typically developing Turkish children ^24^. Children in our cohort attended CWC since approximately three weeks post-earthquake and many continue to do so for routine appointments and camps. A systematic review on peer support for children has detailed how camps can improve health-related quality of life and social abilities and provide a safe environment to discuss their experiences made them feel empowered ^26^. Children in our cohort are known to be in continual contact with each other via social media, maintaining close friendships even if they live in different areas. We expect all this contributed to the excellent social scores for these children.

PedsQL-Emotional scores of children in all amputation groups of our cohort were far lower than those reported by typically developing Turkish children ^24^ and Turkish children with non-earthquake related limb loss ^11^. The cause of this is likely multifaceted but represents the biggest difference between this cohort and children with non-earthquake related limb loss. Emotional challenges could arise from the personal earthquake trauma of the child, such as loss, helplessness and fear ^27^. Seventy-five percent of children lost a first-degree family member and 28.6% lost both their parents. Many children had earthquake-related psychiatric diagnoses. Phantom limb pain prevalence was low compared to adult literature ^28^, but it has a known connection to detrimental emotional pathways ^29^. One child with bilateral lower limb loss reported constant phantom limb pain, which could be related to the especially low emotional scores in that group. Whilst provision of high-quality prosthetics and rehabilitation from government and international donors is important, children who sustain limb loss in a natural disaster, clearly would benefit from early and widespread mental health support.

Emotional challenges in children with limb loss following an earthquake may also result from the influence of their caregiver’s emotions and the turbulence of family units who may themselves be adjusting to loss, anger, helplessness, and hopelessness for themselves and their children ^27^. Furthermore, caregivers will be affected by loss of family who have died or themselves sustained physical or mental trauma and be attempting to secure housing and work amid a disaster zone. Most children in our cohort were living below the local poverty line, with many below the hunger line, and many were still living in emergency shelter accommodation. As might be expected, a high caregiver burden was found in the same cohort of children and their families, and caregiver mental health was associated with depression and anxiety diagnoses in their children ^30^.

## Limitations

Limitations of this study include the retrospective nature of the data, which resulted in small pieces of missing data. Data were collected from children when they attended the CWC for routine appointments. Children may no longer attend the CWC if it is far from their home or if they are now seen by adult services. Two groups were very small (unilateral UL and LL and unilateral UL with bilateral LL), so statistical comparisons were not performed for them. Whilst other groups were small in the broad context of research, they demonstrate a particularly large cohort of children where outcome data are sparse.

## Conclusions

Children with limb loss sustained in the 2023 Kahramanmaraş-Türkiye Earthquakes have good physical, social and school health-related quality of life two years post-amputation. These outcomes are testament to their prosthetic provision, specialist rehabilitation, and treatment within a supportive, social community. Emotional quality of life was much lower in these children due to the personal trauma brought about by the earthquakes and the psychosocial effects of death, injury, and displacement experienced within their family units. Provision of early and widespread mental health support for children and their families after a natural disaster may improve outcomes for future children.

At the time of writing, a devastating earthquake hit Venezuela. Our findings are timely and important and can provide recommendations for care. Understanding long-term outcomes of these children would help plan physical and psychosocial health responses in future natural disasters.

## Data Availability

Data in the present study is not available

## Supplementary Tables

**Supplementary Table SA:** A Amputation level and prosthetic componentry for children in each amputation group. (SACH: solid ankle, cushion heel foot component)

| Group | Amputation |  | Prosthetic |  |  |  |
|---|---|---|---|---|---|---|
|  | Level | n | Suspension | Knee | Terminal device | n |
| **Unilateral upper limb** | Shoulder disarticulation | 1 | - | - | 6-channel | 1 |
|  | Transhumeral | 10 | - | - | 6-channel | 1 |
|  |  |  | - | - | 4-channel | 7 |
|  |  |  | - | - | Cosmetic | 2 |
|  | Transradial | 2 | - | - | 2-channel | 1 |
|  |  |  | - | - | Cosmetic | 1 |
|  | Partial hand/finger | 2 | - | - | Cosmetic | 1 |
|  |  |  | - | - | None | 1 |
| **Unilateral lower limb** | Hip disarticulation | 1 | Belt system | Fixed/mechanical | SACH | 1 |
|  | Transfemoral | 5 | Modular | Hydraulic | Carbon | 2 |
|  |  |  | Pin lock | Hydraulic | Carbon | 1 |
|  |  |  | Pin lock | Fixed/mechanical | Dynamic | 1 |
|  |  |  | KISS | Hydraulic | Dynamic | 1 |
|  | Knee disarticulation | 8 | Classic | Fixed/mechanical | SACH | 1 |
|  |  |  | Modular | Hydraulic | Dynamic | 1 |
|  |  |  | Pin lock | Hydraulic | Dynamic | 1 |
|  |  |  | Pin lock | Hydraulic | Carbon | 3 |
|  |  |  | Passive vacuum | Fixed/mechanical | Dynamic | 1 |
|  |  |  | Passive vacuum | Pneumatic | Dynamic | 1 |
|  | Transtibial | 13 | Modular | - | Dynamic | 3 |
|  |  |  | Modular | - | Carbon | 2 |
|  |  |  | Modular | - | SACH | 2 |
|  |  |  | Pin lock | - | Carbon | 2 |
|  |  |  | Pin lock | - | Dynamic | 1 |
|  |  |  | Passive vacuum | - | Carbon | 2 |
|  | Syme | 1 | Modular | - | Carbon | 1 |
|  | Chopart | 1 | Modular | - | Carbon | 1 |
|  | Partial foot/toe | 2 | Classic | - | Cosmetic | 1 |
|  |  |  | None | - | - | 1 |
| **Bilateral lower limb** | Hip disarticulation + transfemoral | 1 | Belt system + modular | Fixed/mechanical + Fixed/mechanical | Dynamic + dynamic | 1 |
|  | Transfemoral + transfemoral | 2 | Pin lock + pin lock | Hydraulic + hydraulic | Carbon + carbon | 2 |
|  | Knee disarticulation + transtibial | 1 | Passive vacuum + pin lock | Pneumatic | Carbon + carbon | 1 |
|  | Transtibial + transtibial | 2 | Modular + modular | - | Carbon + carbon | 1 |
|  |  |  | Classic + classic | - | Dynamic + dynamic | 1 |
|  | Transtibial + partial foot/toe | 1 | Passive vacuum + none | - | Carbon + none | 1 |
| **Unilateral upper and unilateral lower limb** | Transfemoral + partial hand/finger | 1 | Pin lock | Hydraulic | Carbon + none | 1 |
| **Unilateral upper and bilateral lower limb** | Shoulder disarticulation + transfemoral + partial foot | 1 | Modular | Fixed/mechanical | Dynamic + cosmetic | 1 |
|  | Transhumeral + transfemoral + transtibial | 1 | Pin lock + modular | Hydraulic | Carbon + carbon + 4-channel | 1 |

